# Positioning zoonotic disease research in forced migration: a systematic literature review of theoretical frameworks and approaches

**DOI:** 10.1101/2021.03.19.21253943

**Authors:** Alex Tasker, Dorien Braam

**Affiliations:** Department of Anthropology, University College London, United Kingdom; Disease Dynamics Unit, University of Cambridge, United Kingdom

**Author notes:** Corresponding author (AT). Both authors contributed equally to this work.

## Abstract

**Background:** The emergence and transmission of zoonotic diseases are driven by complex interactions between health, environmental, and socio-political systems. Human movement is considered a significant and increasing factor in these processes, yet forced migration remains an understudied area of zoonotic research – due in part to the complexity of conducting interdisciplinary research in these settings.

**Objectives:** We conducted a systematic review to identify and analyze theoretical frameworks and approaches used to study linkages between forced migration and zoonotic diseases.

**Methods:** We searched within eight electronic databases: ProQuest, SCOPUS, Web of Science, PubMed, PLoSOne, Science Direct, JSTOR, and Google Scholar, to identify a) research articles focusing on zoonoses considering forced migrants in their study populations, and b) forced migration literature which engaged with zoonotic disease. Both authors conducted a full-text review, evaluating the quality of literature reviews and primary data using the Critical Appraisal Skills Programme (CASP) model, while theoretical papers were evaluated for quality using a theory synthesis adapted from Bonell and Fletcher (2013). Qualitative data were synthesized thematically according to the method suggested by Noblit and Hare (1988).

**Results:** Analyses of the 23 included articles showed the increasing use of interdisciplinary frameworks and approaches over time, the majority of which stemmed from political ecology. Approaches such as EcoHealth and One Health were increasingly popular, but were more often linked to program implementation and development than broader contextual research. The majority of research failed to acknowledge the heterogeneity of migrant populations, lacked contextual depth, and insufficient acknowledgement of migrant agency in responding to zoonotic threats.

**Conclusions:** Addressing the emergence and spread of zoonoses in forced migration contexts requires more careful consideration and use of interdisciplinary research to integrate the contributions of social and natural science approaches. Robust interdisciplinary theoretical frameworks are an important step for better understanding the complex health, environment, and socio-political drivers of zoonotic diseases in forced migration. Lessons can be learned from the application of these approaches in other hard-to-reach or seldom-heard populations.

## Introduction

Zoonotic diseases represent a growing threat to public health [1], with over 60% of newly emerging human pathogens originating in animal species [2]. This burden is felt unequally across populations, with endemic zoonotic diseases accounting for an estimated 20% of human illness and death in low- and middle-income settings (LMIS) [3]. Despite geographic variations in zoonotic disease impacts, recent pandemic outbreaks of Zika, Ebola, and SARS-CoV-2 provide stark reminders of the global importance of interspecies disease transmission. Zoonotic disease emergence is influenced by intersecting political, economic and social factors, operating from global to local levels [4, 5]. Population movements pose significant challenges to the control and prevention of zoonotic disease [6], yet the impact of migration on infectious disease remains poorly understood [7]. Migration, and forced migration in particular, pose unique theoretical [8], methodological [9], and ethical [10] challenges to zoonotic research as forced migrant populations often sit awkwardly between geographies and research disciplines, resulting in the common treatment of forced migrants as homogenized sub-populations in environmental and political studies [11].

Across disciplines, scholars are increasingly recognising the importance of engaging with forced migrants as a distinct population. At the end of 2019, there were an estimated 79.5 million forced migrants, including 20.4 million refugees, 45.7 million internally displaced persons (IDPs), and 4.2 million asylum seekers [12]. LMIS bear the brunt of these displacements; political instability and disasters drive forced migrants to seek refuge within their own, or neighboring countries. Increased international connectivity has resulted in forced migration with profound impacts on global health [13]; factors such as exposure to new ecological environments and endemic diseases [14], limited sanitation [15], deteriorating health services [16], and increased human- and animal stress [17] are thought to drive the emergence and spread of infectious diseases.

The inherent relational complexities of forced migration and zoonotic diseases require a nuanced understanding of social, biological, and ecological factors which have not been adequately studied to date [18]. Traditional epidemiological approaches may struggle to represent the intersections of biological and social processes, creating barriers to accurate representation of the complex factors driving disease emergence, transmission and distribution [19]. The emergence of biosocial theoretical frameworks from epidemiological approaches have provided alternative tools to account for social interactions for understanding these complex interactions, influenced by their epistemological foundations (see, for example, Kingsley and Taylor (20)). As an important emerging field of study, research into zoonoses and forced migration requires a well-developed understanding of the contexts and drivers of forced migration, along with critical reflexivity of theoretical and methodological biases. This systematic review aims to make a significant contribution to the under-researched topic of zoonoses in forced migration by identifying existing theoretical frameworks and approaches used, and critically apprising these to inform future enquiry.

### Materials and Methods

### Design

Engaging with the multi-layered and intersecting drivers of zoonotic disease in forced migration contexts presents significant challenges for researchers. Evaluating the quality and utility of existing approaches used to study these complex dynamics is a vital first step in expanding our understanding of this understudied area. Our systematic literature review contributes to this emerging knowledge base by answering the research question: how have theoretical frameworks and approaches been utilized to study the role of forced migration in zoonotic disease dynamics?

We designed a systematic literature review to identify, collate, and comparatively analyze qualifying literature on zoonoses in forced migration to explore the theoretical and methodological foundations used in these articles. The original protocol for this study is registered with the Open Science Framework, DOI 10.17605/OSF.IO/E9JUF. The study follows the statements on the Preferred Reporting Items for Systematic Review and Meta-Analysis (PRISMA); the PRISMA flow chart is given in figure 1, the PRISMA statement for this study is provided in supplementary materials S1.

### Search strategy

Eight electronic databases were interrogated in June 2020: ProQuest, SCOPUS, Web of Science, PubMed, PLoSOne, Science Direct, JStor, and Google Scholar, using the search terms zoono* AND (“forced migra*” OR “internal* displace*” OR refugee*). Search terms were developed from wider analyses of zoonotic research, forced migration literature, expert suggestion, and informed by AT and DBs experience. No limit was placed on publication dates.

### Eligibility criteria

Primary inclusion criteria were:

1. Publication of **full-text peer reviewed articles** available in **English**.

2. A **central focus on zoonotic disease**

3. The **specific inclusion** of one or more **forced migrant populations**

4. The **explicit use** of **theoretical frameworks or approaches** at any stage of the study.

### Screening and quality assessment

Citations were downloaded and manually screened for duplicates by the authors (AT and DB); the remaining full articles were reviewed using the inclusion criteria above. Disagreements between the authors were resolved by consensus through discussion. The quality of literature reviews and primary data studies were verified using the standardized Critical Appraisal Skills Program (CASP) [21] approach to systematically assess the validity, rigor and contribution of sources to our research question and objective. There remains a lack of a standardized protocol to assess the quality of theory synthesis [22]; we therefore evaluated the quality of theoretical evidence using criteria drawn from a theory synthesis conducted by Bonell, Fletcher (23). The quality assessment of sources using CASP criteria showed that included literature reviews and primary data were of high quality; the theoretical review revealed a limited number of assumptions, none of which led to disqualification from the study. The results of the combined quality assessment did not result in the exclusion of any studies, but increased the collective rigor of the synthesis [24]. The theoretical quality review is presented in S2, and the CASP quality review in S3.

### Data extraction and synthesis

The synthesis was informed by an inductive qualitative comparative review to identify emergent key themes across the texts in line with the methodology detailed by Noblit and Hare (25). These themes were then interpreted drawing on wider forced migration and development literatures to explore the applications and limitations of each approach, creating six second order themes and two third-order themes, *population characterization and theory and practice*.

## Results

### Search outcomes

The systematic review identified 2,153 articles; removal of duplicates left 2,067 articles. Titles and abstract screening excluded a further 2,004 articles, leaving 63 articles for full-text analysis (see figure 1). Twenty-three articles met the criteria for inclusion and were included in the comparative analysis, these are given in S4.

### Characteristics of included studies

The majority of included publications were theoretical contributions (n = 10), with a limited number of literature reviews (n = 5 and commentary pieces (n = 3). The remaining articles combined meeting reports (n = 2), a lecture (n = 1), prevalence survey (n = 1), and response plan (n = 1). The characteristics of the papers are included in supplementary materials S4.

### Qualitative Analysis

We conducted an inductive qualitative comparative analysis of the articles in line with the methodology described by Noblit and Hare (25). Six second order themes were identified: endemic versus exotic diseases, movement as a vector, homogenisations and generalisations, agency and action, environments and environmental change, health systems and sanitation. These themes were interpreted to identify two second-order themes, population characterization, and theory and practice. The following section reviews the data under these organizing themes.

#### A. Population characterization

Human displacement is driven by a range of complex factors. Within the reviewed literature we identified five typologies of displacement and eight causes, detailed in supplementary materials S5. The majority of articles identified displaced populations as refugees (n = 16), however of these 16, half (n = 8) did not consider the drivers of displacement. Where discussed, conflict (n = 7) and environmental change (n = 5) were the most common drivers; there was a progression towards a more substantiated discussion of conflict and environmental drivers in more recent articles.

Geographically, most publications focused on LMIS populations; 9 from a generalized global view and 11 from a geographically-bounded perspective, including regional (e.g. sub-Saharan or Mediterranean) and country-specific foci (e.g. Turkey or Uganda); details of geographical foci are provided in supplementary materials S6. Despite many studies employing disaster narratives in their framings of specific humanitarian emergencies, there was limited evidence of clustering of locations around discrete disaster events linked to forced migration.

Geographical considerations are central to understanding zoonoses in forced migration. Vulnerabilities of forced migrant populations are rooted in historical and political contexts; drawing on case studies in Africa, Kalipeni and Oppong (26) argue that forced migration should be situated in historical migration events, showing how movements are interlinked with resource scarcity and conflict. The authors note how colonial powers’ common disregard for local contextual understandings has led to the homogenisation of ‘Africa’; the authors cite blind-spots to the uneven distribution of specific refugee populations across the continent as a consequence and symptom of reductionist approaches. Despite these observations, the authors themselves stop short of linking homogenisations to differential individual- and collective disease risks within displaced populations.

The specific contribution of forced migration to zoonotic risks were less well explored. Multiple authors argued that resettlement programs and refugee migration may act as vectors for endemic disease into naive populations, citing increased disease risk through environmental and socioeconomic change such as poverty and health inequality [27-29]. The earliest work included in this review [30] suggested these factors be addressed though improved co-ordination of development programs; while subsequent work by Mayer (31) developed the need to disentangle disease risks in forced migration by improving understanding of biological drivers of disease dynamics. The author argued this was particularly true of dense and unhygienic settlements – forced migrants fell into this category. Despite acknowledging a need for greater depth of understanding, the majority of authors stopped short of engaging with the wider complexities of human migratory movements. Meanwhile, forced migration studies (as opposed to those specifically dealing with zoonoses) recognize how drivers of migration may have significant implications for health [32]. For example, McMichael (33) distinguishes between voluntary and forced migration, noting *that “well-managed migration could provide an adaptive response that reduces the adverse health impacts of climate change”* [33, p.549].

#### B. Theory and practice

The second theme identified tensions between research framed as more abstract theoretical enquiry, and more empirical, holistic approaches informed by a combination of theoretical precedent and contextual understandings. Adherence to structured theory during research remains a contested subject across disciplines, with views ranging from ‘there is nothing so practical as a good theory’ [34, p.18] to suggestions that the use of theory in research may inhibit intellectual engagements [35]. Theoretical foundations, and associated conceptual frameworks, are commonly used to construct and validate both question and evidence, and as such are highly political processes that shape the outcomes of scholarship. We identified the use of fifteen discrete theoretical frameworks across the 23 studies, conducted by researchers from 11 disciplines and sub-disciplines. The first publication to explicitly employ a theoretical framework was published in 1988, with articles steadily increasing over time up to 3 per year in 2015, 2017, and 2018. We observed a subjective movement from the use of social science perspectives such as political ecology (n = 2) and systems theory (n = 2) from the late 1990s-early 2000s towards more cross-disciplinary engagement employing Planetary Health (n = 3) and One Health (n = 7) lenses in the last six years. Our review included four studies (structured reviews (n = 3) and reports (n =1)) which gave little consideration to nuanced discussions around theory. The following section discusses each in turn; firstly more abstract theoretical studies, followed by more contextually-rooted work.

### Disciplines and theoretical frameworks

Kalipeni and Oppong (26) explicitly employed a political ecological framework to suggest how disease dynamics frameworks should move beyond separate consideration of biophysical and environmental factors in forced migration. The authors highlighted how political and social forces underpin refugee crises, leading the authors to conceptualise unsanitary refugee camps as spaces which could lead to the emergence of zoonotic diseases. This framing links political and ecological drivers of refugee movement to negative health outcomes for forced migrants, through which the authors explicitly advocated for high-level responses to protect global health against zoonotic disease. These responses, they argue, must be understood through a nuanced multilevel historical contextualization of intersections between environmental and societal factors, including human agency in environmental changes.

Themes of environmental change can be found in many of the included studies. Writing two years later, Mayer (31) again used a political ecological approach to explore disease ecologies of emerging zoonotic infections, this time rooted in a biomedical framework which incorporated the unintended consequences of human actions from individual to global levels. Pedersen (36) developed the political aspects of environmental change further, using a human rights-based lens to argue that ecosystems are overburdened by the ’pursuit of unlimited economic growth’ [36, p.745]. This view placed the responsibility of zoonotic mitigation on high-level policy makers to directly engage with disease emergence. Pedersen argues in their study that natural resource conflict and development synergistically drive forced migration, which results in the burdens of disease being borne by the displaced; an insightful argument that resonates with many contemporary investigations of links between migration and health inequalities [37]. Despite a promising theme of enquiry, the author does not develop this position further to suggest how specific impacts of disease dynamics are driven, or felt by forced migrant populations.

Parallel to macro-political landscapes of inequalities and health, many studies in this review structured their research so as to present empirical case studies which explored the influence of national and international health systems on disease pathways. Spatial diffusion theory used by Bouma and Rowland (38) characterized Pakistani refugee camps as disease transmission nodes, driven by overcrowding and unsanitary living conditions, but limited attention was given to the specific mechanisms by which this may occur. An alternative exploration of disease diffusion was given by Schærström (39) who emphasized the individual and environmental challenges posed by these sites, once again the mechanics of these factors was left uncovered. In contrast, Singer (40) explicitly highlighted anthropogenic factors in mass population migration which may contribute to zoonoses through the development of syndemic theory. In their earlier paper, Singer suggested how microbial changes are linked to social and environmental conditions which in turn underpin zoonotic threats; these themes are revisited in their later paper through consideration of social justice, disease prevalence, and health inequalities driven by poverty, stress, and structural violence [41]. We suggest that this later article represents a step forward in bridging multiple dimensions of zoonotic disease emergence in forced migration, most notably through the recognition of heterogeneity within specific study sites at global and local levels.

As noted earlier, social justice was discussed by both Pedersen (36) and Singer (40). In their report on the Anthropocene to The Rockefeller Foundation and Lancet Commission, Whitmee, Haines (42) continue this debate using a Planetary Health framework, calling for the integration of ’multisectoral actors’ [42, p.2006] to transform health systems. A Planetary Health framework is again employed by Myers (43) to explore case studies of Ebola, HIV, and schistosomiasis through which they highlight links between human-ecosystem relationships and disease exposure risks. Myers argues that environmental changes are driven by social processes and population characteristics; this position is echoed by Flowra and Asaduzzaman (29) who consider climatic drivers of microbes and infection as linked to social changes. In parallel to the growth in non-forced migration literature exploring intersections between social and global environmental change, many of the later papers in this review recognized climatic change as a key factor in understanding the emergence and transmission of zoonoses. Myers (43) suggested environmental changes drive negative health outcomes for forced migrants through processes of malnutrition, infectious disease, and trauma. In parallel, Tomlinson, Hunt (44) detail the role of global environmental change as a potential driver of forced migration. Both of these studies frame their discussion of zoonotic disease outbreaks as a feature of increased overcrowding, deforestation and wildlife encroachment; both studies once again stop short of fully exploring the complexity and variability of the social and biological mechanisms leading to these outcomes.

#### Approaches for research and implementation

When evaluating zoonoses in forced migration, it is important to understand the often opaque relationship between theoretical and applied approaches used by researchers. For example, political ecology is often characterized as a theoretical approach, whereas frameworks including One Health and EcoHealth resist easy definition, variously employed as theory, methodology, and a policy devices [45] – tools for both research and implementation. One Health approaches in this study were most commonly described as frameworks for either a) exploring eco-social processes driving infectious disease emergence and spread in human and animal populations [27], or b) tools for program development [28]. In this review, One Health approaches were specifically applied to the behavior and movement of reservoir species for zoonotic disease pathogens [46, 47], and the exploration of zoonotic disease risks from poverty and poor hygiene [48]. These studies varied in location, population, and depth, ranging from granular explorations of respiratory infections in Kenyan and Thai refugee settlements [28] and Ebola in Congolese refugees in Ugandan camps [49], to more macro-level evaluations of Rift Valley Fever [46] and Nipah [50]. In this last example, the total engagement with One Health was as an aim for future work. Regardless of the topic and depth, no study which employed One Health acknowledged the role of historical, political and socio-economic drivers in their evaluations.

EcoHealth approaches in forced migration research emerged almost in parallel with One Health. EcoHealth shares much common ground with One Health, providing systems-based ways of thinking around zoonoses, disease emergence, and pandemic threats [51]. EcoHealth is often cited as contributing a more nuanced understanding of ecosystem drivers of disease than One Health [52], though it is unhelpful to consider either as a superior tool for holistic evaluation [53]. In the studies considered for this review, EcoHealth approaches were most commonly used to explore land use change and disease; using the framework, Patz, Daszak (54) discuss refugee movements as vectors for Tuberculosis and Hepatitis B, Confalonieri and Aparicio Effen (55) suggest Nipah links to land use change and Lyme disease through increased contact with host diversity, and Kittinger, Coontz (56) explore interactions with snail hosts of *Schistosomiasis*.

## Discussion

Despite covering over three decades of work, the articles considered in the review consistently display a number of shortcomings. Most articles treated forced migrants as exemplar populations, proxies for exposure to specific and isolated risks, without acknowledgement or consideration of the inherent complexity and diversity of these populations. Multiple studies reviewed here chose to link human mobility to endemic and non-endemic disease exposure, reducing forced migrants to vectors for the movement of non-endemic disease into study populations. We argue that the reality for forced migrants is significantly more complex. Classifying populations as forced migrants for instance overlooks the fact that many within these communities may have been settled for many years [30]; indeed evidence suggests that disease patterns are similar between long-established forced migrant populations and host populations living in comparable living conditions [38]. From Africa [57], Asia [58], to Europe [59] and beyond, researchers are increasingly arguing for robust consideration of endogenous heterogeneity of migrant populations, and those at the margins of state control. For example both newly migrated, and long-term residents are situated within widely differentiated social networks [60]. These relationships offer diverse and differentiated resources that can shape zoonoses transmission, detection, and treatment. This complexity is compounded by the highly contextual nature of many endemic zoonotic diseases, operating within distinct ecological ranges. We argue that this renders generalisations solely on the basis of national borders unhelpful, especially when considered through the lens of long-term informal cross-border movements. From our previous research, we support the idea that communities labeled as *migratory* may often have more in common with settled indigenous groups in border zones than the wider national population. This underlines the importance of understanding political and social context when framing zoonotic disease risks in migrant populations, in particular the unique institutional landscape in which many refugees exist [61].

In addition to internal diversity and contextual detail, all studies considered in this review failed to consider migrant-led responses to zoonotic disease. Many articles cited poor sanitation and ineffective health systems, yet forced migrants have in the past adapted their surroundings through local water supply innovations [62], and developed internal networks for health provision [63]. In the last decade, multiple studies have detailed the endogenous creativity of migrant populations [64], to the point that refugee self-reliance has on occasion been co-opted as a development tool, arguably to the detriment of the communities in question [65, 66]. Despite external political influence, without acknowledgment of migrants’ individual agency, and the dynamic nature of forced migrants’ relationships with zoonotic disease, we suggest that research is likely to fail to accurately capture the complexity of these sites of disease emergence and transmission.

Our review does give cause for hope. The subjective shift from the political ecological approaches of the 1990s and early 2000s, towards a more Public Health-based understanding in the last decade suggests a movement towards a more holistic, interdisciplinary treatment of complexity. These approaches are arguably better positioned to map the nonlinear nature of zoonoses which are driven by a variety of social, political, historical, economic, environmental, and biological factors. Advances in One Health and EcoHealth approaches have arguably outpaced wider understandings of the role of human migration on infectious disease, but we are optimistic that increasing attention offers the chance to bring these domains together. Despite this optimism, the centering of Public Health in forced migration zoonotic research should prompt critical reflection, informed by current debates on neocolonial aspects of international aid and development programming. Public Health has the ability to generate highly techno- and ethnocentric narratives which, rather than engaging local communities in healthcare, can serve to obfuscate indigenous knowledge systems and further disadvantage the most vulnerable. Considerations of social justice can be found across articles include in the review [31, 36, 44]; as research moves to engage with the inherent complexity of zoonotic research in forced migration, we suggest that actors involved in forced migration, including national- and international NGOs, governments, and activist groups, should explore how these approaches may reflect local understandings and systems of power, rather than providing a façade for the business-as-usual of international assistance.

### Limitations of the review

There are several limitations to this review. Only English language articles were included, limiting our engagement with indigenous scholarship. Secondly, it is possible that ethnographic studies exist that engage with zoonotic diseases using locally-defined terms which would not have been detected by the search protocol. This review only considered peer reviewed articles, making publication biases likely and selecting against internal reports from development organizations. Given the focus of this review was primarily theoretical, it reports and evaluations which did not explicitly employ theoretical or analytical frameworks will have been excluded.

## Conclusions

The results of this review indicate that research on zoonotic diseases in forced migration increasingly employs interdisciplinary approaches to consider holistically social, environmental, and biological complexity. These approaches acknowledge linkages between human, animal, and environmental health through the integration of disciplines and participation of local stakeholders and interlocutors. Many of these approaches build on political ecological approaches to address macro-level disease risks, but often overlook the complexity and diversity of forced migrant populations. Studies continue to characterize movement as a major determinant for disease and health outcomes, despite a lack of evidence for doing so. Disease vulnerabilities depend on a multitude of internal and external factors which must be better researched and contextualized; further work is required to build theoretical frameworks able to address the societal and biological processes of forced migration across time and space.

## Supporting information

Supplementary Table 6 displacement typologies

Supplementary Table 5 Publication characteristics

Supplementary Table 3 Theoretical quality review

Supplementary Table 4 CASP quality review

Supplementary Figure 1 PRISMA flow diagram

## Data Availability

Systematic Review protocol detailing the the manuscript

## Supplementary materials

S1 Table: PRISMA Statement

S2 Table: Theoretical quality review

S3 Table: CASP Quality Review

S4 Table: Publication characteristics and inclusion grid

S5 Table: Typologies and causes of displacement

S6 Table: Geographical focus and distribution of studies

