## Supplementary Table 6 displacement typologies for "Positioning zoonotic disease research in forced migration: a systematic literature review of theoretical frameworks and approaches"

### S5 Table: Typologies and causes of displacement

|  | **Article number (see S4)** | | | | | | | | | | | | | | | | | | | | | | |
| --- | --- | --- | --- | --- | --- | --- | --- | --- | --- | --- | --- | --- | --- | --- | --- | --- | --- | --- | --- | --- | --- | --- | --- |
|  | **1** | **2** | **3** | **4** | **5** | **6** | **7** | **8** | **9** | **10** | **11** | **12** | **13** | **14** | **15** | **16** | **17** | **18** | **19** | **20** | **21** | **22** | **23** |
| ***Displacement typology*** | | | | | | | | | | | | | | | | | | | | | | | |
| Resettle | x |  | x |  |  |  |  |  |  |  |  |  |  |  |  |  |  |  |  |  |  |  |  |
| Refugee |  | x |  |  | x | x | x | x |  | x |  | x | x | x |  | x |  | x | x | x | x | x | x |
| IDP |  |  |  |  |  |  |  |  | x |  |  |  |  |  | x |  |  |  |  |  |  |  | x |
| Migrant |  |  |  | x |  |  |  |  | x |  |  |  |  |  |  |  |  | x |  |  |  |  |  |
| Forced Migration |  |  |  |  |  |  |  |  |  |  | x | x |  |  |  |  |  |  |  |  |  |  |  |
| Unspecified |  |  |  |  |  |  |  |  |  |  |  |  |  |  |  |  | x |  |  |  |  |  |  |
| ***Displacement Driver*** | | | | | | | | | | | | | | | | | | | | | | | |
| Drought | x |  |  |  |  |  |  |  |  |  |  |  |  |  |  |  |  |  |  |  |  |  |  |
| Rural-urban | x |  |  |  |  |  |  |  |  |  |  |  |  |  |  |  |  |  |  |  |  |  |  |
| Returnee | x |  | x |  |  |  |  |  |  |  |  |  |  |  |  |  |  |  |  |  |  |  |  |
| Conflict |  | x |  |  |  |  |  |  |  |  |  |  |  |  | x |  | x |  | x | x | x | x |  |
| Unspecified |  |  |  | x | x | x | x | x |  |  | x |  |  | x |  | x |  | x |  |  |  |  |  |
| Development |  |  |  |  |  | x |  |  | x |  |  |  |  |  |  |  |  |  |  |  |  |  |  |
| Environmental change |  |  |  |  |  |  |  |  |  | x |  |  | x |  | x |  | x |  |  |  |  |  | x |
| Conservation |  |  |  |  |  |  |  |  |  |  |  | x |  |  |  |  |  |  |  |  |  |  |  |
