## Supplementary Table 5 Publication characteristics for "Positioning zoonotic disease research in forced migration: a systematic literature review of theoretical frameworks and approaches"

### S4 Table: Publication characteristics and inclusion grid

| **Item** | **Author, Year** | **Affiliation** | **Publication field** | **Type of study** | **Location** | **Displacement** | **Effect on zoonoses** | **Theory/ approach** |
| --- | --- | --- | --- | --- | --- | --- | --- | --- |
| 1 | Kloos et al., 1998 [30] | Geography | Social Science | Literature review | Ethiopia | Resettlement, refugee, pastoralism, labour migration | Disease distribution | Three-factor disease complex |
| 2 | Bouma and Rowland, 1995 [38] | Medicine | Medicine | Prevalence survey | Pakistan | Refugees | Not direct | Sota-mogi model |
| 3 | Aiken et al., 1996 [67] | Army | Environment | Review | Bosnia-Herzegovina | Refugees | Importance to military personnel | Environmental health |
| 4 | Pedersen, 1996 [36] | Medicine | Social Science | Literature review | Global | Shifting settlement patterns | Interactions human health, development and environmental change | Undefined framework |
| 5 | Kalipeni and Oppong, 1998 [26] | Geography | Social Science | Review | Africa | Refugees | Causes of forced migration influence health risks | Political ecology |
| 6 | Mayer, 2000 [31] | Geography | Social Science | Review | Global | Refugees, development displaced | Environmental changes | Political ecology |
| 7 | Patz et al., 2004 [54] | Environmental health | Environment | Meeting report | Global | Refugees | Environmental and land use change | Systems model approach |
| 8 | Schærström, 2009 [39] | Geography | Geography | Review | Global | Refugees | Disease distribution | Disease diffusion |
| 9 | Kittinger et al., 2009 [56] | EcoHealth | Ecology and health | Literature review | China | Development displaced | Change in social-ecological dynamics | EcoHealth |
| 10 | Confalonieri and Effen, 2011 [55] | Science and Technology | Environmental health | Literature review | Global | Environmental refugees | Human encroachment on ecosystems | EcoHealth |
| 11 | Singer, 2014 [40] | Medical Anth | Anthropology | Review | Global | Forced migration | Environmental change and overpopulation | Syndemics |
| 12 | Nicole, 2014 [68] | Conservation | Environmental health | Commentary | Uganda | Conservation refugees | Increased poaching | One Health, EcoHealth |
| 13 | Machalaba et al. | Global Health | Global Health | Literature review | Global | Climate change refugees | Expanded suitable habitat pathogens and vectors | One Health, systems approach |
| 14 | Nanyingi et al., 2015 [69] | Medicine | Biomedical science | Literature review | Sub-saharan Africa and Arabian peninsula | Refugees | Disease distribution | One Health |
| 15 | Whitmee et al., 2015 [42] | Environmental health | Medicine | Commentary | Global | Internal displacement, conflict | Effect environmental change on disease distribution | Planetary health |
| 16 | Jones et al., 2017 [27] | Veterinary Medicine | Medicine | Review | Global | Refugees | Ecosocial processes, land-use change | One Health |
| 17 | Myers, 2017 [43] | Environmental health | Medicine | Lecture | Global | Conflict and environmental displacement | Human-mediated ecosystem effects | Planetary Health |
| 18 | Singer et al., 2017 [41] | Anthropology | Medicine | Review | Global | Refugees | Health inequality | Syndemics, biosocial complex |
| 19 | Inci et al., 2018 [70] | Veterinary Medicine | Medicine | Review | Turkey | Refugees | Socioeconomic and environmental changes | One Health |
| 20 | Flowra and Azadussaman, 2018 [29] | Public Health | Planetary Health | Commentary | Bangladesh | Refugees | Socioeconomic: overcrowding and hygiene | Planetary Health |
| 21 | Abubakar et al., 2018 [28] | Public Health | Public Health | Meeting report | Kenya and Thailand | Refugees | Assumed | One Health |
| 22 | Aceng et al., 2020 [49] | Global Health | Public Health | Preparedness plan review | Uganda | Refugees | Social drivers of disease spread | One Health |
| 23 | Tomlinson et al., 2020 [44] | Medicine | Global Health | Review | Global | Climate change displacement | Intersections between social, economic, and environmental drivers around mental health | Planetary Health |
