## Supplementary Table 3 Theoretical quality review for "Positioning zoonotic disease research in forced migration: a systematic literature review of theoretical frameworks and approaches"

### S2 Table: Theoretical quality review

| **Authors** | **Research design** | **Evidence theoretical approach** |  |  | **Theoretical engagement** |  |
| --- | --- | --- | --- | --- | --- | --- |
|  | *Theoretical approach* | *Constructs specified* | *Causal pathways* | *Suggestions* | *Limitations/ applicable* | *Assumptions* |
| Aiken et al., 1996 [67] | Environmental health | Historical, political background; military implementation; medical and public health overview; military health issues; data collection system; veterinary health | Interrupted infrastructure and services supply affecting health; social issues affecting health | Interdisciplinary responses; environmental health responses limit non battle injuries; need for inclusion of veterinary public health | Lack of application in non-conflict setting; military health responders limits sustainability; need to ; monitor animal health status | Limited theorization of environmental health |
| Pedersen, 1996 [36] | Ecological model of health | Comprehensive disease ecology; conventional epidemiological model, inorganic, organic and cultural components; expanded with ecosystem, climate change, social, economic and political systems; based in ethics of economic growth, conflict, and nationalism | Disease is a result of interaction of the body with disruptive environmental inputs; development impacting the environment and through this mental, physical and social health | A new disease ecology needs to be translated into a new research agenda involving ecologically-oriented health and social scientists; enlarge the framework of analysis | To enhance models to show relationship between variables for hypothesis generation, testing and refinement; taking into account historical, political and social context | Contradictory and unsupported claim that 'some vectors and zoonoses' might have been eliminated through land clearance |
| Kalipeni and Oppong, 1998 [26] | Political ecology | Political / social forces interact with biophysical or adverse environmental conditions to produce disease; geographies of exile and refugee movements and the associated implications for re-emerging and newly emerging infectious diseases | Circumstances underlying the refugee crisis influence health service delivery and disease in camps; increasing globalization puts the world at risk of infections | Proposed solution is mainly based on high-level political reform, not multilevel | The approach is useful for refugee and displacement situations in its multilevel approach, and inclusion of historical analysis | Includes little biological analysis, instead focusing on the disruption of health services and fertile disease environment, as well as behaviour |
| Mayer, 2000 [31] | Political ecology | Context and scale refer to multi-scale contextual analysis; combines disease ecology with concepts of political economy, acknowledging both human-made and natural components, to explain dynamics | Human-environment relations only understood by analysing relationships of patterns of resource use; centrality human agency and structure (institutions restricting actions of agent) | Political ecology can alter the concepts of the causality of disease from a purely biomedical concept to one that also incorporates the unintended aspects of human action | Society-environment interactions require a historical analysis | Does not provide an extensive review and critique of the theory |
| Patz et al., 2004 [54] | Systems model approach | Identifies complexity of land use change and the risks and benefits to human health | Anthropogenic land use changes drive a range of infectious disease outbreaks and emergence events and modify the transmission of endemic infections | Policy-relevant levels of the model include specific health risk factors, landscape or habitat change, and institutional (economic and behavioural) | What if displacement does not cause land use change | Lack of inclusion of endemic disease |
| Schærström, 2009 [39] | Spatial diffusion theory | Concepts and methods from epidemiology, geography and cartography useful to understand, describe and analyse the processes leading to new spatial patterns of ill health | Infections will spread with human and animal movement; changing distribution of non-infectious diseases; character of the process depends on the disease and geographically different physical and social circumstances | With increasing mobility and environmental change, disease diffusion is an urgent public health issue on local, regional, national, and global levels | When considering changing geographical distribution and intensity of diseases | Need for further methodological and theoretical development |
| Confalonieri and Effen, 2011 [55] | Ecohealth/ Ecosystem concept/ approach | Eco-epidemiology paradigm, conceptual frameworks for environmental health studies include ecosystem characteristics and changes as determinants of health | Deteriorating environmental conditions are a major contributory factor to poor health and poor quality of life | Interdisciplinary collaboration of epidemiologists, ecologists, environmental scientists, and social scientists required | Changes in natural systems in a global scale are important; drivers of human health is also a challenge for public health research and practice | Article focuses on environmental/ ecosystem changes |
| Singer, 2014 [40] | Syndemics | Zoonotic infections pose syndemic threats; microbe's inherent properties and opportunities by their evolutionary trajectories; by social and environmental conditions | Anthropogenic factors contributing to zoonoses: climate change, deforestation, overpopulation, disruptions due to military actions, mass migrations of populations due to disasters, inadequate food and water supplies | The product of multiple species behaviours and interspecies interactions at all levels: multispecies ethnography brings a holistic lens | Need to consider how many species to include, etc. | Lacks practical implementation approach to identify drivers beyond concepts |
| Nicole, 2014 [68] | One Health; Ostrom's economic theory; Ecohealth | Connects humans, animals and ecosystems; conservationists and healthcare professionals to improve community health and livelihoods | Conservation refugees; evicting increases poaching [and disease] | One Health shows much promise; projects must explicitly account for the political, social, and economic settings; need interdisciplinary thinking and collaboration: | Markets and states often failed to protect both ecosystems and human livelihoods; give the local people most invested in using a common resource a say in its management to solve this social–ecological dilemma | Lacks theoretical depth, consideration and use of concepts |
| Whitmee et al., 2015 [42] | Planetary health | Planetary health; environmental changes; multifactor actors; investments | Internal displacement, conflict/ environmental displaced: impact on health; effects of global environmental change on the spread of zoonoses | Planetary health calls for improved health systems, with sufficient attention to environmental health, the integration of multisectoral actors, and balancing investments between the health of present and future populations | Primarily considering animal health within food security | Economic model not specific to zoonoses |
| Jones et al., 2017 [27] | One Health/ ecosocial processes | One Health; Eco-social processes inﬂuencing infectious disease emergence and spread | Provides examples of emerging infectious disease to illustrate eco-social processes | One Health approach used to better understand complexity and connectedness of eco-social processes on the emergence and spread of infectious diseases amongst humans and animals | Need to embrace systems-thinking and inter-disciplinary approaches | Improved understanding of epidemiological and eco-social processes, including their interdependence, is essential |
| Myers, 2017 [43] | Planetary health | Description of the scale of human impacts on natural systems and the extensive associated health effects | Disease ecology research shows anthropogenic change affecting the risk of exposure to infectious diseases; vulnerability to a particular set of biophysical changes in the environment is determined by characteristics of the population itself | In the context of planetary health, the boundaries between public health and nearly every other facet of human activity become more porous; new paradigm requires a new science to address research priorities | Researchers and public health practitioners to work across disciplines and embrace partnerships | Need data on forced displacement effect on human health: malnutrition, epidemic infectious disease, physical, sexual, and psychological trauma, and mental illness |
| Singer, 2017 [41] | Syndemics | Syndemics model: biosocial complex of interacting, co-present, or sequential diseases and the health inequality | Social and environmental factors that promote and enhance the negative effects of disease interaction; syndemics most likely to occur in places of health inequality caused by poverty, stigmatisation, stress, or structural violence | Structural interventions within the biomedical setting can have a greater impact than conventional clinical interventions on disease control | Based in medical anthropology, biosocial concept of syndemics offers a holistic approach to address synergistic disease and context interactions | More inclusive than One Health approach, what if non-clustering disease |
| Flowra and Asaduzzaman, 2018 [29] | One Health and Planetary Health | One Health; Planetary health; synergistic; migratory and vulnerable populations | Climate-health nexus model shows multiple relationships with climate variability (linear negative effect of temperature and positive effect of overcrowding and immigration on the incidence of infection | Need for interdisciplinary prevention; need for a migratory and vulnerable population health information repository, that includes public and animal health and environmental statistics | Planetary health and One Health to work together as an interdisciplinary collaboration | Assumption that ‘livestock is well known risk' |
| Abubakar et al., 2020 [28] | One Health | One Health; epidemiological and virological surveillance, outbreak detection and response,  influenza at the animal-human interface | Need to track bird migratory routes to prevent influenza outbreaks | One health approach is essential for cross-species surveillance of pandemic influenzas | One Health framework for action was drafted to enhance surveillance and better data collection at the human-animal interface | no direct link refugees and zoonoses; assumptions |
| Aceng et al., 2020 [49] | One Health | One Health; to implement preparedness, surveillance and screening programme | Uganda’s experience in Ebola virus disease outbreak preparedness, 2018–2019 | This approach requires coordination across multiple sectors of government including human and animal health, agriculture, wildlife, water and environment, security, immigration and law enforcement | VD plan was also developed based on the One Health approach to Global Health Security at both national and subnational levels | Need to sustain efforts across spectrum of multi-hazard framework |
| Tomlinson et al., 2020 [44] | Planetary health; Social, economic and environmental determinants of global mental health | Planetary health to think systemically about the interactions between planet, climate, and human systems | Climate related change can strongly influence or exacerbate conflict with the consequent catastrophic impact on migration and population mental health; climate breakdown, overcrowding, deforestation, and encroachment into wildlife territories contribute to outbreaks of zoonotic diseases | New innovative political and economic approaches are essential on order to protect the planet’s natural systems, and thereby the health and development of humans for generations | Capacity to manage zoonotic disease outbreaks will be increasingly challenged by increasing population movement, urbanization, and fragile health systems | Less research on the potential impact of the increasing risk of infectious diseases on mental health |
