## Supplementary Table 4 CASP quality review for "Positioning zoonotic disease research in forced migration: a systematic literature review of theoretical frameworks and approaches"

### S4 Table: CASP Quality review

**CASP: Literature Review**

| **Authors** |  | **CASP SR** |  |  |  |  |  |  |
| --- | --- | --- | --- | --- | --- | --- | --- | --- |
|  |  | **Are the results valid (A)** | |  |  |  | **What are the results (B)** | |
|  | Type of study | Focused question | Right type of papers incl? | All relevant studies included? | Assessed quality of studies? | Combined results | Overall results | How precise results |
| Kloos et al., 1988 [30] | Literature review | YES | YES | YES | YES | YES | high confidence | high confidence |
| Kittinger et al., 2009 [56] | review | YES | YES | YES | CAN'T TELL | YES | high confidence | high confidence |
| Machalaba at al., 2015 [47] | Literature review | YES | YES | YES | CAN'T TELL | YES | moderate confidence | CAN'T TELL |
| Nanyingi et al., 2015 [69] | literature review | YES | YES | YES | CAN'T TELL | YES | high confidence | high confidence |
| İnci et al., 2018 [70] | review | YES | YES | YES | CAN'T TELL | YES | high confidence | high confidence |

**CASP: Empirical Study**

| **Authors** |  | **CASP** |  |  |  |  |  |  |  |  |
| --- | --- | --- | --- | --- | --- | --- | --- | --- | --- | --- |
|  |  | **Are the results valid (A)** | |  |  |  |  | **What are the results (B)** | |  |
|  | *Type of study* | *Focused* | *Cohort recruitment* | *Exposure* | *Outcome* | *Confounding factors* | *Follow up* | *Results* | *Precise* | *Believe* |
| Bouma, and Rowland, 1995 [38] | prevalence survey | YES | YES | YES | highly confident | YES | CAN'T TELL | model confirmed | YES | YES |
