## Supplementary Figure 1 PRISMA flow diagram for "Positioning zoonotic disease research in forced migration: a systematic literature review of theoretical frameworks and approaches"

### S1 PRISMA flow diagram

Full-text articles excluded, with reasons

(n = 40)

Lack of theoretical detail

(n = 28)

Lack of displacement detail (n = 10)

Non-zoonotic disease

(n=2)

Records excluded
(n = 2,004 )

Records screened for evaluation of title and abstract
(n = 2,067)

Full-text articles assessed for eligibility
(n = 63)

Studies included in qualitative synthesis
(n = 23)

Records after duplicates removed
(n = 2,067)

Additional records identified through expert inclusion
(n = 6)

Records identified through database searching
(n = 2,147)

### Included

### Eligibility

### Identification

### Screening
